# Early genomic detection of SARS-CoV-2 P.1 variant in Northeast Brazil

**DOI:** 10.1101/2021.02.25.21252490

**Authors:** Stephane Tosta, Marta Giovanetti, Vanessa Brandão Nardy, Luciana Reboredo de Oliveira da Silva, Marcela Kelly Astete Gómez, Jaqueline Gomes Lima, Cristiane Wanderley Cardoso, Tarcisio Oliveira Silva, Marcia São Pedro Leal de Souza, Pedro H. Presta Dia, Vagner Fonseca, José Lourenço, Luiz Carlos Junior Alcantara, Felicidade Pereira, Arabela Leal

**Affiliations:** Laboratório de Genética Celular e Molecular, Instituto de Ciências Biológicas, Universidade Federal de Minas Gerais, Belo Horizonte, Minas Gerais, Brazil; Laboratório de Flavivírus, Instituto Oswaldo Cruz, Fundação Oswaldo Cruz, Rio de Janeiro, Brazil; Coordenação Geral dos Laboratórios de Saúde Pública/Secretaria de Vigilância em Saúde, Ministério da Saúde, (CGLAB/SVS-MS) Brasília, Distrito Federal, Brazil; KwaZulu-Natal Research Innovation and Sequencing Platform (KRISP), School of Laboratory Medicine and Medical Sciences, College of Health Sciences, University of KwaZulu-Natal, Durban, South Africa Department of Zoology; Centro e Informações Estratégicas de Vigilância em Saúde do estado da Bahia (CIEVS); Secretaria Municipal de Saúde de Salvador; Diretoria de Vigilância Epidemiológica do Estado da Bahia (DIVEP); Secretaria Municipal de Saúde de Irecê; University of Oxford, Oxford OX1 3PS, UK; Laboratório Central de Saúde Pública da Bahia – LACEN-BA, Salvador, Bahia, Brazil

**Keywords:** SARS-CoV-2, P.1 variant, Genomic surveillance, Brazil

## Abstract

Tracking the spread of SARS-CoV-2 variants of concern is crucial to inform public health efforts and control the ongoing pandemic. Here, we report genetic evidence for circulation of the P.1 variant in Northeast Brazil. We advocate for increased active surveillance to ensure adequate control of this variant throughout the country.

**Article Summary Line:** Active genomic surveillance of SARS- CoV-2 suspected cases from recent travelers reveals the circulation of the P1 variant of concern in Bahia state, Northeast Brazil.

## Text

Since the emergence of the severe acute respiratory syndrome coronavirus-2 (SARS-CoV-2) in 2019, the combination between the unprecedented number of cases and more than 500k genomes generated has allowed the identification of hundreds of circulating genetic variants during the pandemic *(1)*. Currently, three variants (B.1.1.7 or VOC202012/01, B.1.351 or 20H/501Y.V2 and P.1) carrying several mutations in the receptor-binding domain (RBD) of the spike (S) protein, raise concerns about their potential to shift the dynamics and public health impact of the pandemic *(2-5)*. They appear potentially associated with (i) increased transmissibility, (ii) propensity for re-infection, (iii) escape from neutralizing antibodies, and (iv) increased affinity for the human ACE2 receptor *(6-8)*.

First identified in January 2021 in travelers from the Amazonas state (North of Brazil) who arrived in Japan, the P.1 variant (alias of B.1.1.28.1) *(9)*, harbors a constellation of 17 unique mutations, including three in the receptor binding domain of the spike protein (K417T, E484K, and N501Y). It thus immediately raised concerns to public health authorities over the risk of its unknown potential of faster spreading and/or worsening of coronavirus disease (COVID-19) clinical outcomes. In view of its rapid spread in Brazil and elsewhere *(4,5,10)* the public health authorities of the Bahia state (Northeast Brazil) conducted an active monitoring for a rapid detection of this variant in the state.

Here, we report genetic evidence of the circulation of the P.1 variant in Bahia, by generating 11 SARS-CoV-2 complete genomes from travelers returning from the Amazonas state (North of Brazil). In mid-January 2021, routine genomic surveillance in the Central Laboratory of Health of the Bahia state, started a massive screening of COVID-19 patients and their contacts reporting a travel history to/from the Amazonas state, resulting in eleven suspected SARS-CoV-2, P.1 infections. Viral RNA was extracted from nasopharyngeal swabs and tested for SARS-CoV-2 by multiplex real-time PCR using SARS-CoV-2 assay Allplex^™^ 2019-nCoV Assay (Seegene) and Charité: SARS-CoV2 (E/RP) (Biomanguinhos). Cycle threshold values (Cts) for the common target between the two assays, gene E, ranged from ≈16 to ≈35. We next subjected the qRT-PCR–positive samples to viral genomic amplification and sequencing using the Ion GeneStudio^™^ S5 Plus Ion Torrent (Life Technologies, USA), according to the manufacturer’s instructions. Consensus sequences from the eleven positive tested samples were generated by *de novo* assembling using Genome Detective (https://www.genomedetective.com/) *(11)*. A total of 13,234,814 mapped reads were obtained, resulting in a sequencing mean depth of 4824 and a coverage of 98%. The new whole genome sequences generated where assigned, according to the Pangolin *(1)* lineage classification, as the recently identified P.1 variant (alias of B.1.1.28.1). All patients had recently record travel from the city of Manaus in Amazonas back into the Bahia state. Four patients were from the same family and the other others had no other known connection between them. The data from the eleven patients are described in **(Table 1)**.

**Table 1.**
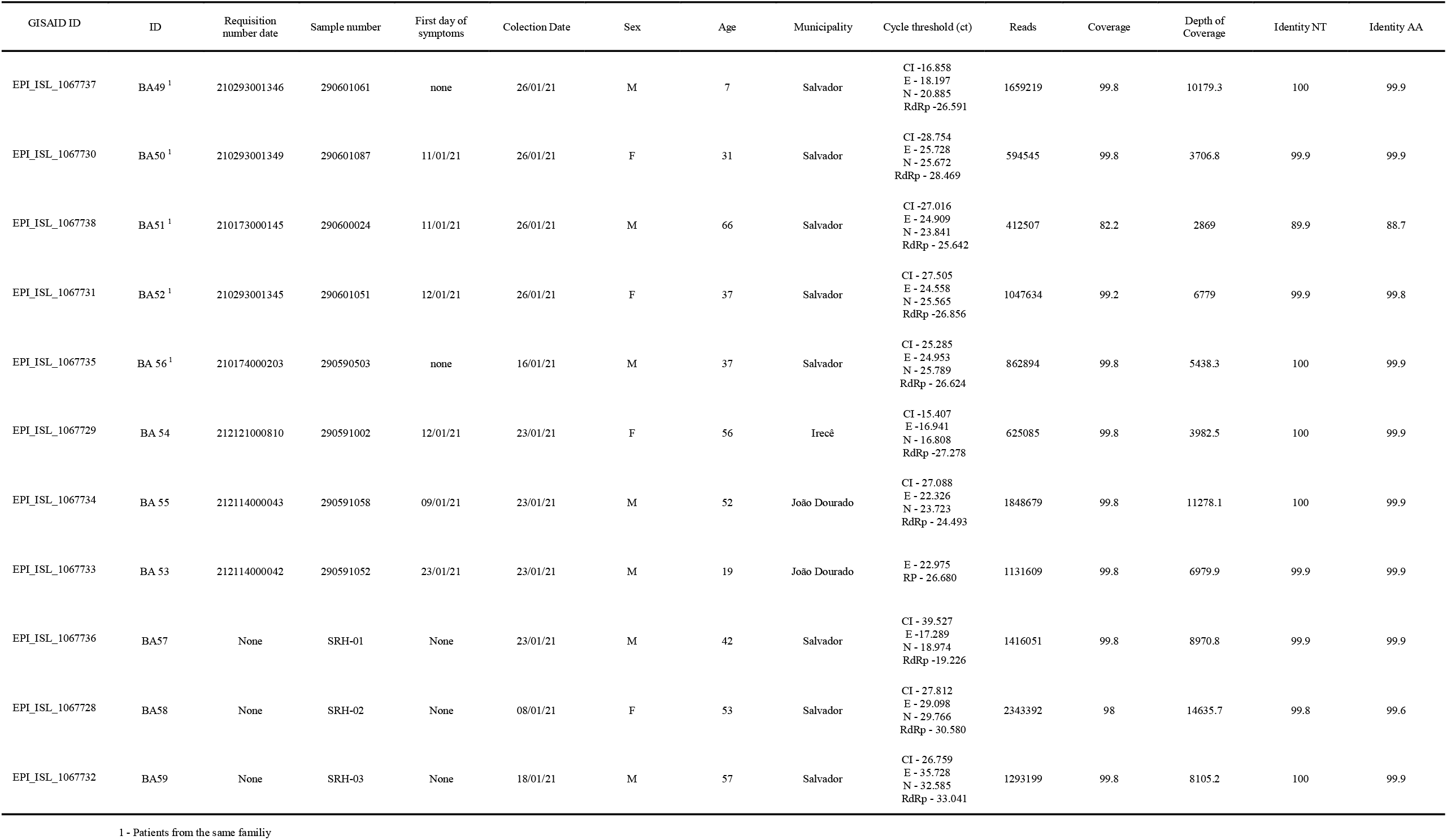
Information from patients with travel history from Manaus, Amazonas to state of Bahia.

We explored the genetic relationship of the newly sequenced P.1 genomes to those of other isolates by phylogenetic inference. To do so, we combined the eleven new isolates (Accession numbers EPI_ISL_1067728 - EPI_ISL_1067738) with all Brazilian SARS-CoV-2 (n=1663) genomes, including recently released P.1 genomes (**Figure 1 panels A-B**) *(4,5)* available on GISAID (https://www.gisaid.org/) up to February 21st, 2021. Only genomes >29,000bp and <1% of ambiguities were considered (n=1663). Sequences were aligned using MAFFT *(12)* and submitted to IQ-TREE for maximum likelihood (ML) phylogenetic analysis *(13)*.

**Figure 1.**
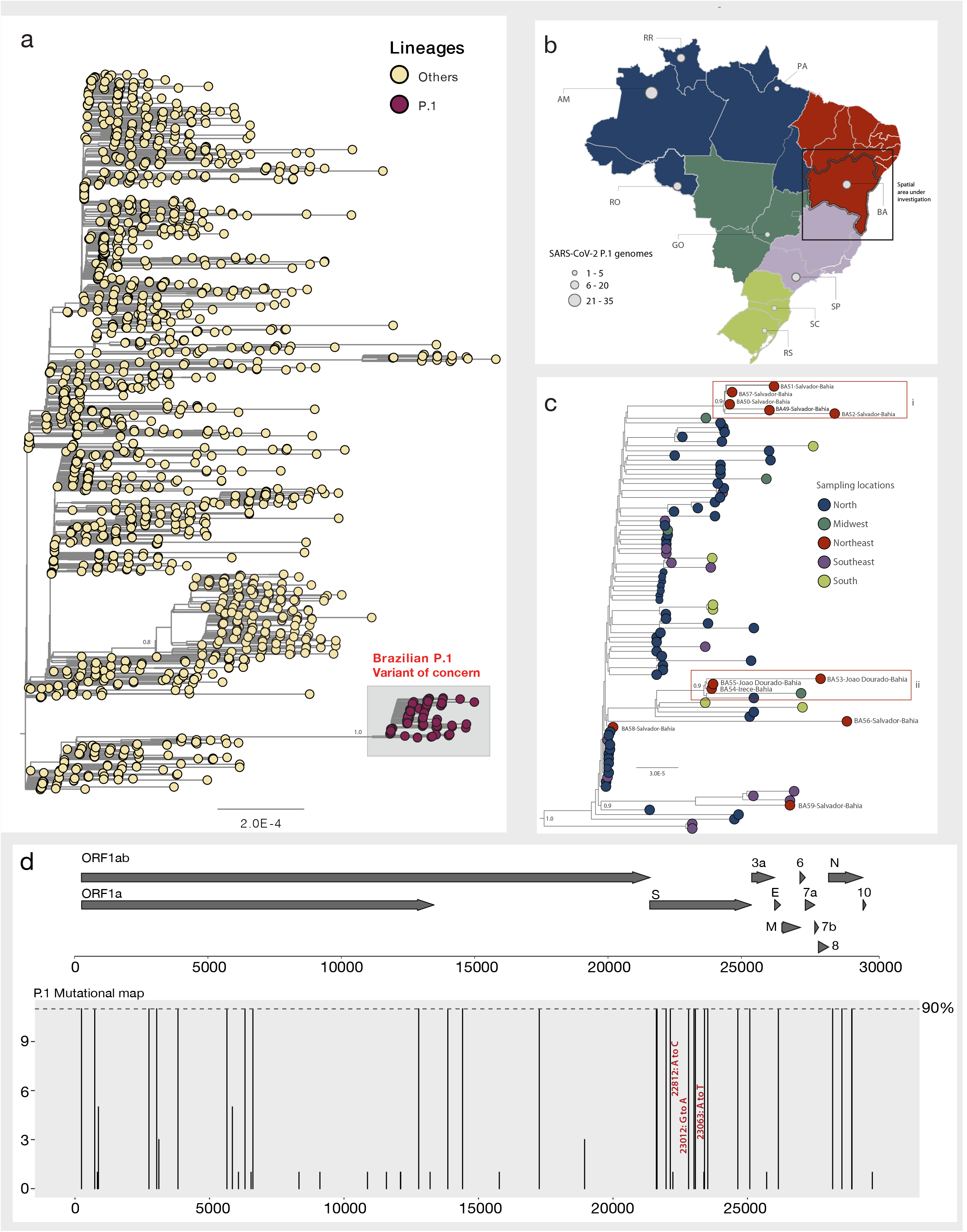
Genomic detection of the SARS-CoV-2 P.1 variant of concern in Bahia state, Northeast Brazil. a) Maximum likelihood (ML) phylogenetic tree including the 11 new isolates obtained in this study plus n=1663 SARS-CoV-2 Brazilian strains collected up to February 21st, 2021. b) Map of Brazil showing the number of P.1 SARS-CoV-2 sequences available by region and state. North region (blue): AM=Amazonas state, RR=Roraima state and RO= Rondônia state; Midwest region (dark green): GO=Goiás state; Southeast region (lilac): SP=São Paulo and South region: SC (light green): Santa Catarina state and RS=Rio Grande do Sul state and Northeast region (red): BA=Bahia state. c) Representation of the zoom of the Brazilian P.1 clade. Branch support (SH-aLTR >0.8) is shown at key nodes. d) Variant maps of the P.1 lineage-defining-mutations were mapped against the SARS-CoV-2 genome structure. Most common mutations defined as mutations present in more than 90% of the genomes in that group. Mutations of international concern: i) K417T (22812A>C); ii) E484K (23012G>A) and iii) N501Y (23063A>T), among the RDB domain are highlighted in red.

Phylogenetic estimation strongly supported placement of the isolates from Bahia within the Brazilian P.1 clade (**Figure 1, panel A**) (SH-aLTR = 1.0). Our results further highlighted that most of our new isolates formed two well supported monophyletic clusters (**Figure 1 panel C**). One of those clusters (Cluster i), included samples from the capital city of Salvador, isolated from individuals from the same family of the eleven patients described here (BA49, 50, 51 and 52) plus the sample BA57 (SH-aLTR = 1.0). And a second cluster (Cluster ii) which included samples BA53, 54 and 55 that were isolated from patients from the municipalities of João Dourado and Irecê (SH-aLTR = 1.0). Moreover, samples BA56 and BA58 and BA59 appeared to be interspersed among P.1 strains from the North and Southeast Brazil. By combining all the P.1 strains already available from distinct Brazilian regions (**Figure 1 panel B, C**) our analysis further revealed that this variant was already detected in the majority of Brazilian regions (starting from the North to Midwest, Northeast, Southeast and South Brazil) highlighting the high connectivity of the country and reinforcing the need of active monitoring to follow the local real-time spread of this new variant of international concern. Finally, among the eleven new genomes, we also identified lineage-specific mutations, including the ones of international concern among the RDB domain: K417T (22812A>C), E484K (23012G>A) and the N501Y (23063A>T) (**Figure 1, panel D**).

This report describes the early detection of the SARS-CoV-2 P.1 variant in the Northeast of Brazil (Bahia state) and provides evidence regarding the P.1 circulation across all Brazilian macro-regions that would have occurred within the past two months. The P.1 variant is now known to have emerged in the Amazonas state, but was first detected in travelers arriving in Japan from Brazil. The latter is a testimony for the currently scarce genomic surveillance in Brazil, which has failed to detect the variant before it became a public health emergency in the Amazonas state. Moreover, the eleven individuals here described to carry the P.1 variant in the Bahia state were only detected after a unique active screening initiative that focused on travelers and their recent contacts. This case study should serve as an example of the effectiveness of active surveillance to monitor the importation of genetic variants of importance for public health. Since such initiatives are not only critical to monitor the ongoing spread of known variants but are also necessary to detect the possible emergence of new ones in the near future, continual and revamped investment is needed and will be necessary for adequate public health policy in Brazil.

## Data Availability

Newly generated SARS-CoV-2 sequences have been deposited in GISAID under accession numbers EPI_ISL_1067728 to EPI_ISL_1067738.

## Acknowledgments

This work was financed by Laboratório Central de Saúde Pública da Bahia (LACEN-BA) and Secretaria da Saúde do Estado da Bahia (SESAB) and supported by Pan American Health Organization PAHO/WHO. We also thank Centro e Informações Estratégicas de Vigilância em Saúde (CIEVS) in the municipality of Salvador, Bahia. ST is supported by the Coordenação de Aperfeiçoamento de Pessoal de Nível Superior – Brasil (CAPES) – Finance Code 001. MG is supported by Fundação de Amparo à Pesquisa do Estado do Rio de Janeiro (FAPERJ). JL is supported by a lectureship from the Department of Zoology, University of Oxford. We also would like to thank all the authors who have kindly deposited and shared genome data on GISAID. A table with genome sequence acknowledgments can be found in **Supplementary Table 1**.

## Author Bio

MSc. Fraga is a PhD student at the Federal University of Minas Gerais, Southeast Brazil. Her research focuses on the implementation of next generation sequencing techniques to track the spread of emerging and re-emerging viral pathogens.

## Ethical Statement

This research was approved by the Ethics Review Committee of the Federal University of Minas Gerais (CEP/CAAE: 32912820.6.1001.5149 approval number).

## AUTHOR CONTRIBUTIONS

**Conception and design:** SFT, MG, FPM, and AL; **Data collection:** SFT, MG, FPM, CC, TS, PD, MS; **Investigations:** ST, LS, VN, MKAG, JGL, CC, MG, VF, JL; **Data Analysis:** ST, MG, VF; **Draft Preparation:** ST, MG, JL, FPM, and AL; **Revision:** LCJA, SFT, MG, FPM, CC, TS, PD, MS.

## References

1. O’Toole Á, Scher E, Anthony Underwood, Ben Jackson, Verity Hill, JT McCrone, Chris Ruis, Khali Abu-Dahab, Ben Taylor, Corin Yeats, Louis du Plessis, David Aanensen, Eddie Holmes, Oliver Pybus, Andrew Rambaut. Pangolin: lineage assignment in an emerging pandemic as an epidemiological tool. (github.com/cov-lineages/pangolin)

2. Rambaut A, Loman N, Pybus O, et al. Preliminary genomic characterisation of an emergent SARS-CoV-2 lineage in the UK defined by a novel set of spike mutations. https://virological.org/t/preliminary-genomic-characterisation-of-an-emergent-sars-cov-2-lineage-in-the-uk-defined-by-a-novel-set-of-spike-mutations/563. (Accessed Dec 21, 2020).

3. Tegally H, Wilkinson E, Giovanetti M, et al. Emergence and rapid spread of a new severe acute respiratory syndrome-related coronavirus 2 (SARS-CoV-2) lineage with multiple spike mutations in South Africa. medRxiv 2020.12.21.20248640 (Preprint); doi: https://doi.org/10.1101/2020.12.21.20248640.

4. Faria NR, Claro IM, Candido D, et al. Genomic characterisation of an emergent SARS-CoV-2 lineage in Manaus: preliminary findings. Virological. Date accessed: February 24, 2021. https://virological.org/t/genomic-characterisation-of-an-emergent-sars-cov-2-lineage-in-manaus-preliminary-findings/586

5. Rezende, Paola. “Phylogenetic Relationship of SARS-CoV-2 Sequences from Amazonas with Emerging Brazilian Variants Harboring Mutations E484K and N501Y in the Spike Protein.” Virological, 11 Jan. 2021, virological.org/t/phylogenetic-relationship-of-sars-cov-2-sequences-from-amazonas-with-emerging-brazilian-variants-harboring-mutations-e484k-and-n501y-in-the-spike-protein/585.

6. “Emerging SARS-CoV-2 Variants.” Centers for Disease Control and Prevention, Centers for Disease Control and Prevention, www.cdc.gov/coronavirus/2019-ncov/more/science-and-research/scientific-brief-emerging-variants.html.

7. Vasques Nonaka CK, Franco MM, Gräf T, de Lorenzo Barcia CA, de Ávila Mendonça RN, de Sousa KAF, et al. Genomic evidence of SARS-CoV-2 reinfection involving E484K spike mutation, Brazil. Emerg Infect Dis. 2021 May

8. Weisblum Y, Schmidt F, Zhang F, et al. Escape from neutralizing antibodies by SARS-CoV-2 spike protein variants. Elife 2020; 9:3–10.

9. National Institute of Infectious Diseases, Japan. (2021, January 12.) Brief report: New Variant Strain of SARS-CoV-2 Identified in Travelers from Brazil [Press release]. Retrieved from https://www.niid.go.jp/niid/en/2019-ncov-e/10108-covid19-33-en.htmlexternal

10. Pangolin: lineage assignment in an emerging pandemic as an epidemiological tool (https://cov-lineages.org/global_report_P.1.html)

11. Vilsker, M., Moosa, Y., Nooij, S., Fonseca, V., Ghysens, Y., Dumon, K., … & de Oliveira, T. (2019). Genome Detective: an automated system for virus identification from high-throughput sequencing data. Bioinformatics, 35(5), 871–873.

12. Nakamura, T., Yamada, K. D., Tomii, K. & Katoh, K. Parallelization of MAFFT for large-scale multiple sequence alignments. Bioinformatics. 2018; 34:2490–2492.

13. Nguyen, L.-T., Schmidt, H. A., von Haeseler, A. & Minh, B. Q. IQ-TREE: A Fast and Effective Stochastic Algorithm for Estimating Maximum-Likelihood Phylogenies. Mol Biol Evol. 2015; 32:268–274.

